# Specific and intensive language rehabilitation on post-stroke aphasia using eCALAP: a single-case experimental design

**DOI:** 10.1101/2025.03.25.25324445

**Authors:** Cécilia Jubin, Jeanne Badault, Anne-Catherine Bachoud-Lévi, Marie Villain, Charlotte Jacquemot

## Abstract

**Background and purpose:** Effective rehabilitation needs to be intensive, specific, and suited to the clinical reality of care. Based on previous work, we developed the eCALAP, a tool for aphasia evaluation that generates a tailored rehabilitation program that can be used with the therapist or independently. The aim of this study is to demonstrate the effectiveness of the eCALAP rehabilitation program for people with aphasia (PWA).

**Methods:** Five PWA were included in this concurrent multiple baseline Single-Case Experimental Design (SCED). Repeated measures consisted of a picture naming task and were collected 3 times a week. Baseline duration was randomized and lasted 1 or 2 weeks. Rehabilitation with the eCALAP lasted 3 weeks with 3 rehabilitation sessions per week. For contrasting baseline and rehabilitation performance, statistical analyses were performed according to SCED guidelines. Visual analysis included mean baseline values and a 2-SD envelope post-intervention. Tau-U and Nonoverlap of All Pairs (NAP) analyses were conducted.

**Results:** All PWA were able to complete the entire protocol. The rehabilitation program was effective for four out of the five participants. Tau-U values ranged from 0.44 to 0.72 (p<.05), indicating large effect sizes and strong trends in performance improvement. These results are further supported by the NAP values, which were consistently high, suggesting that the intervention led to substantial changes in the repeated measures as compared to the baseline period.

**Conclusion:** The results of this study provide promising evidence for the effectiveness of the eCALAP rehabilitation program.

## Introduction

Aphasia, an acquired language impairment which can affect both production and comprehension of language, occurs approximately in 40% of stroke cases (Engelter et al., 2006). Aphasia delays the rehabilitation of other cognitive impairments, increases the risk of depression, and reduces the chances of reintegration into active life (Burfein et al., 2024). Aphasia rehabilitation is a cornerstone of post-stroke care but is hindered by clinical and practical challenges. To be efficient, rehabilitation must be intensive and include targeted linguistic tasks specifically designed to activate and restore the affected linguistic components (Bachoud-Lévi et al., 2022; Jacquemot et al., 2012; Shrubsole et al., 2018) The clinical difficulty of setting up a targeted rehabilitation program is now tackled through the eCALAP-evaluation, a battery which selects the most effective tasks according to each patient’s deficits (Bachoud-Lévi et al., 2022). Implementing intensive rehabilitation within the practical realities of clinical care is challenging for several reasons. Many patients encounter barriers to accessing rehabilitation services, such as geographic constraints and transport difficulties. Rehabilitation requires sustained effort over time, but fatigue and lack of motivation often prevent stroke patients from adhering to programs (Bullier et al., 2020; Riley, 2017). Additionally, the limited availability of trained professionals reduces the feasibility of frequent and close rehabilitation sessions.

Digital therapy (DT) offers a promising solution to address these limitations by providing accessible and flexible rehabilitation. DT enables higher therapy dose without increasing the therapist’s workload. To manage patients’ fatigue, sessions can be broken down during the day, while still meeting the required duration. However, despite its popularity, there is limited evidence on the effectiveness of aphasia rehabilitation software (Vaezipour et al., 2020).

One of the primary limitations of current digital therapies is that they often focus on a small range of language impairments, often word retrieval (Cuperus et al., 2024), without addressing the full range of linguistic impairments that vary across patients. Aphasia manifests itself differently in each individual and can affect different language components. As a result, existing digital therapies tend to often adopt a generic approach (Repetto et al., 2020), with exercises that are not targeted or personalized to address each patient’s unique needs thereby diminishing both their effectiveness and medical utility. Most digital therapies have yet to demonstrate robust scientific evidence supporting their efficiency in aphasia rehabilitation. This evidence gap highlights the importance of developing more effective scientifically validated intervention tools. Based on previous work (Bachoud et al. 2022), we therefore developed eCALAP-rehabilitation, a digital tool designed to (1) conduct language evaluation, (2) generate a personalized rehabilitation program, (3) deliver targeted, adaptive linguistic tasks tailored to the patients’ specific impairments. To evaluate intervention efficacy, Single-Case Experimental Designs (SCED) are particularly well suited, as they consider patients’ spontaneous evolution of performance and allow to draw conclusions regarding intervention efficiency with only a small number of patients (Krasny-Pacini & Evans, 2018, Tate et al., 2016). This study aims to demonstrate the effectiveness of the eCALAP rehabilitation program for post-stroke aphasic patients using a SCED methodology.

## Methods and materials

### Participants

Recruitment took place at the Henri Mondor Hospital in Créteil in July 2024. Participants had to present aphasia, confirmed by the CALAP evaluation. All people with aphasia (PWA) were right-handed and French speakers. Participants were excluded when illiterate, presented uncorrected hearing and/or visual impairments, or if they had a known neurodegenerative disorder. Five people with aphasia (PWA) were enrolled in this study, all of whom experienced aphasia following a single event of left hemisphere ischemic stroke (see Supplementary for demographic table). Four of them had chronic aphasia (>3 months post-stroke) while one was in the sub-acute phase (<3 months post-stroke). Aphasia ranged from mild to severe on the ASRS scale, but all presented sufficient comprehension to understand the main goal of the study and the linguistics tasks.

### Experimental design

This study was approved by the institutional ethics review board (N°IRB: 00012024-71; N° IDRCB: 2022-A00489-34), and all patients provided written consent to participate. This study had a concurrent multiple baseline design across participants, sequencing the onset of the intervention.

The study consisted of two phases: baseline (A) and rehabilitation (B). During both phases, PWA performance was measured repeatedly using a picture-naming task provided at a constant frequency throughout the entire protocol, with sessions held three times a week. All sessions were conducted by the same therapist.

The duration of the baseline was randomized between 4 and 6 sessions using a Research Randomizer program (http://www.randomizer.org), to cap the duration of the study and minimize patient dropout. During the baseline sessions, picture naming tasks were administered, but no rehabilitation was provided to the participants. Language assessment using the eCALAP was conducted at the end of baseline to generate a tailored rehabilitation program. The rehabilitation phase lasted 9 sessions, all participants received 3 sessions of rehabilitation per week, during which the therapist conducted the repeated measures and the rehabilitation program, using the eCALAP-rehabilitation program. A tablet was provided to the participants at the start of the intervention phase enabling them to do self-rehabilitation sessions (SRS) between appointments with the speech therapist, and therefore increasing the rehabilitation dose.

### Material

The repeated measure consisted of a picture naming task. Fifteen lists of 30 items were constructed. Word frequency was matched between lists. Word order was randomized across PWA.

The eCALAP-rehabilitation program consists of a list of rehabilitation tasks that are performed digitally using the eCALAP application on a Samsung Tablet (SM-T575). Linguistic tasks within the program were selected from a list of 62 including comprehension, production, and repetition tasks. Each task was decomposed into the different components it activates within the language model of Figure 1a. In the eCALAP-rehabilitation program, the training tasks that were selected were the ones specifically targeting the impaired language components (for details, see Bachoud et al., 2022). Each task offered multiple levels of difficulty so they could be tailored to each participant and aphasia severity.

**Fig. 1:**
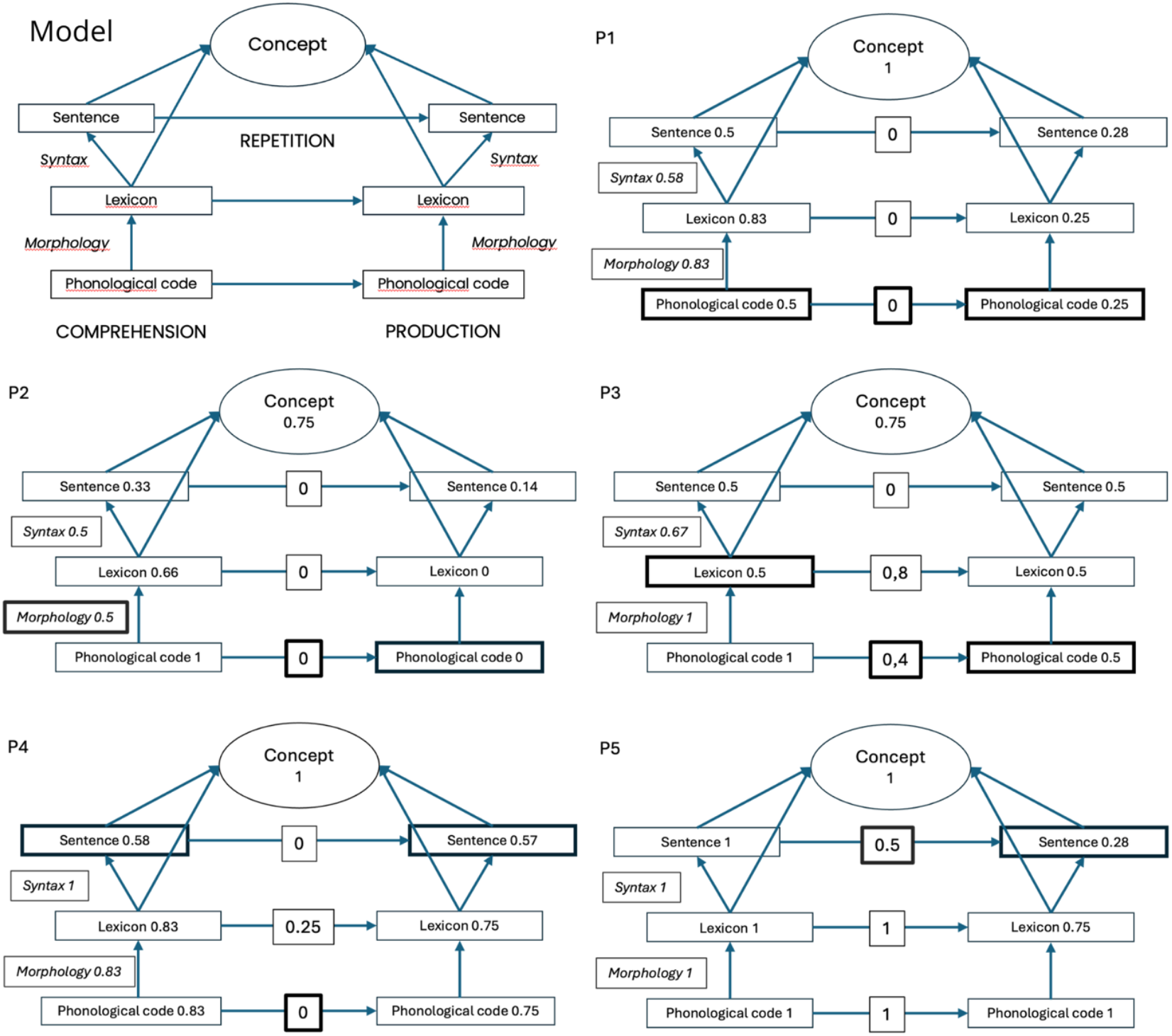
Patients’ linguistic profiles according to the CALAP model. Psycholinguistic model of oral language processing (upper left) with Core Assessment of Language Processing (CALAP). The model distinguishes the comprehension, production, and repetition modality and within each modality, the different components: phonology (set of speech sounds), morphology (how the sounds combine to form words), lexicon (words), syntax (how words combine to form sentences) and concept (semantic knowledge) that can be selectively impaired. The CALAP allows for evaluating each component of this model. The scores of the linguistic components were normalized according to each participant’s performance, so the component’s best score was attributed a value of 1 and the lowest score a value of 0. The normalization enables the identification of the components the most impaired for each participant with the lowest value. Bold components represent the components that were selected for the rehabilitation program. The rehabilitation program selected the linguistic component the most impaired in each modality (input, repetition and output). If two normalized scores of the same modality were equally impaired, the program selected the smallest linguistic unit for intervention.

### Statistical analysis

For visual analysis, we displayed the mean value during the baseline period along with the 2-SD envelope after the onset of the intervention. Additionally, baseline stability was assessed for each period (i.e., baseline and intervention) using the 80%-20% rule (Manolov & Moeyaert, 2017). Statistical tests were selected in accordance with the SCED guidelines (Krasny-Pacini & Evans, 2018). Visual and statistical analyses were conducted using two online platforms specialized in SCED analysis (Manolov & Solanas, 2018; Tarlow, 2016). To assess picture naming accuracy, we first estimated the monotonic trend of the baseline via Tarlow’s (2016) website. No baseline correction was needed. Tau-U values (Parker et al., 2011) were therefore calculated without baseline correction.

## Results

### Procedural fidelity

Four out of the five participants completed the protocol. P3 missed one rehabilitation session due to a fall. All sessions with the therapist were 45 minutes long.

### Overall effect of the therapy

The rehabilitation program was effective for 4 participants as shown by the picture naming task performances (Fig 2; Table 1). self-rehabilitation sessions (SRS) played a role in increasing the rehabilitation dose. Participants varied in their engagement with the SRS, with the total number of sessions ranging from 2 to 12. The mean duration per session ranged from 3.38 to 14.49 minutes, and the total time spent on self-rehabilitation varied between 16.6 and 136.34 minutes across participants. The number of exercises completed per session was consistent (around 3 exercises per session on average). These individual differences highlight varying levels of adherence and engagement with the digital rehabilitation program. (See Supplementary for detailed SRS data.)

**Table 1:**
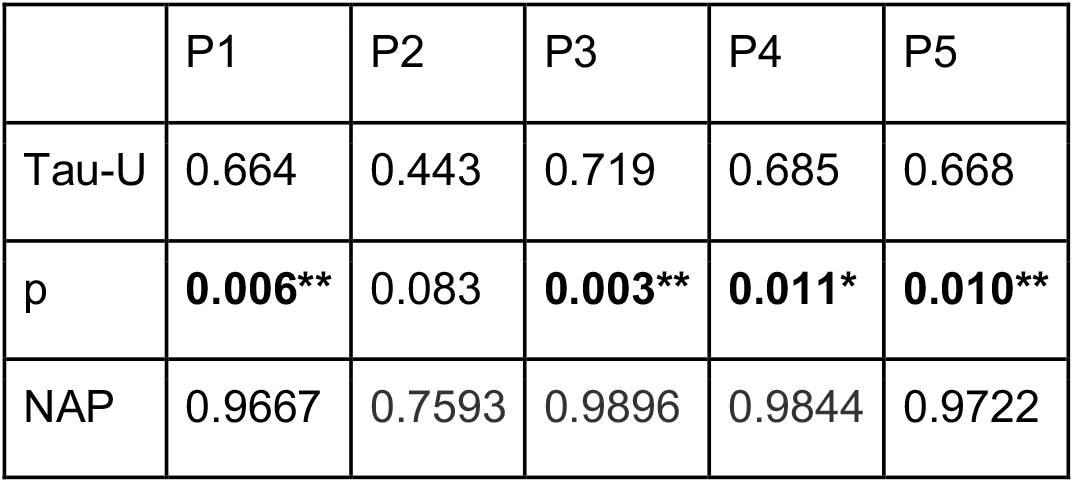
Results of Tau-U analysis and NAP (Nonoverlap of All Pairs) scores for each participant in the eCALAP rehabilitation program. The p-values significativity of the tests are reported with *: 0.01 < p ≤ 0.05, **: 0.001 < p ≤ 0.01.

|  | P1 | P2 | P3 | P4 | P5 |
| --- | --- | --- | --- | --- | --- |
| Tau-U | 0.664 | 0.443 | 0.719 | 0.685 | 0.668 |
| p | **0.006\*\*** | 0.083 | **0.003\*\*** | **0.011\*** | **0.010\*\*** |
| NAP | 0.9667 | 0.7593 | 0.9896 | 0.9844 | 0.9722 |

**Fig 2:**
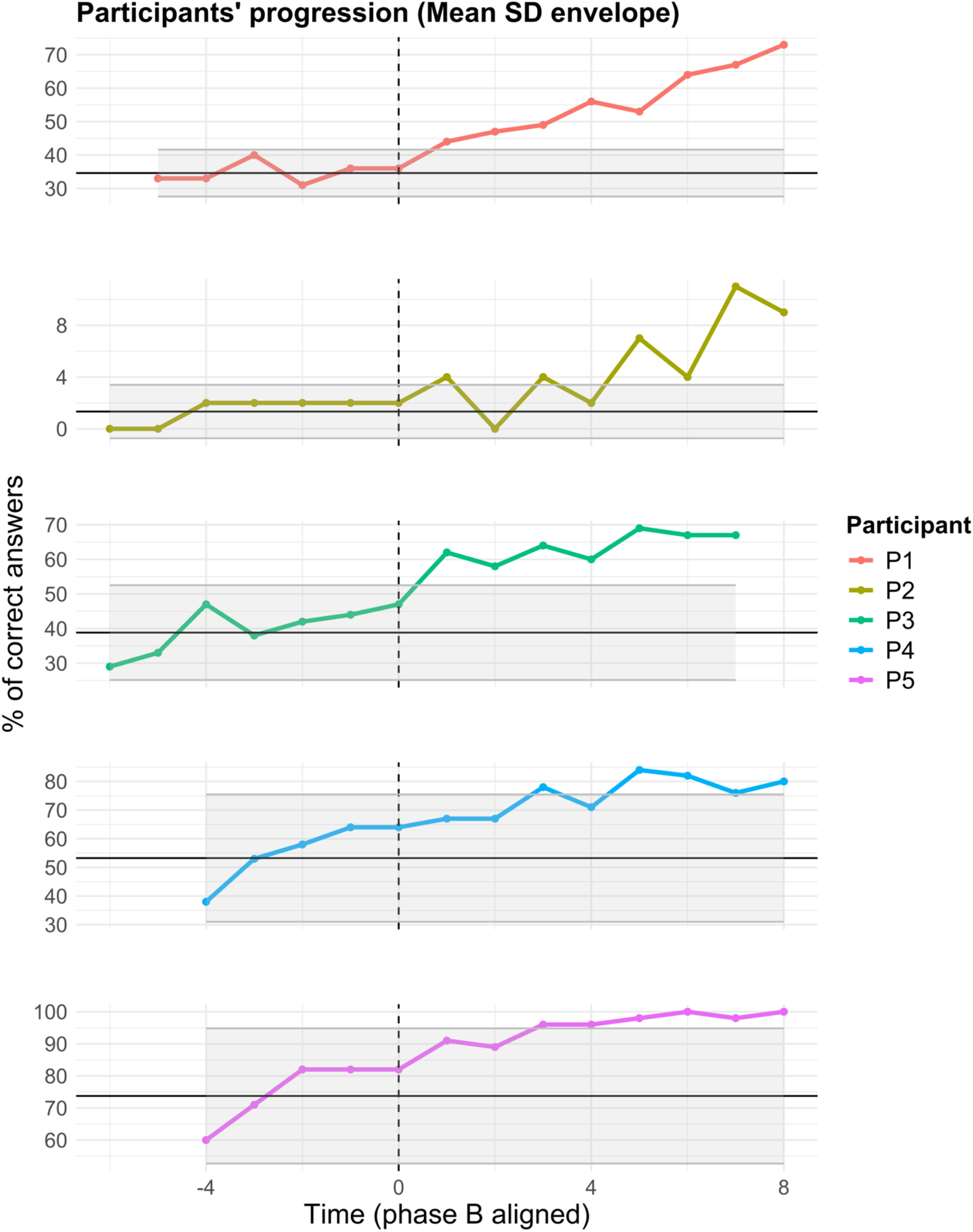
Visual analysis for naming performances of the 5 participants. In grey, we displayed the mean value during the baseline period along with the 2-SD envelope after the intervention started. Session number (3 sessions per week) is displayed on the x-axis. The time 0 indicates the beginning of the intervention phase for each patient. This phase is aligned to improve the visual representation of the results. Naming performance scores are reported on the y-axis.

Tau-U values for these four participants ranged from 0.66 to 0.72, indicating large effect sizes and strong trends in performance improvement. These results are further supported by the NAP (Non-Overlapping Data) values, which were consistently high, suggesting that the intervention led to substantial changes in the picture naming task, as compared to the baseline period. The p-values for these participants (all < 0.05) also confirm the efficacy of the rehabilitation program. P2’s progression however was more mild than the other participants’ but NAP value (0.75) still demonstrated moderate effect.

## Discussion

The results of this study provide promising evidence for the effectiveness of the eCALAP-rehabilitation program in aphasia therapy. The positive effects observed in the picture naming task, particularly for four out of the five participants, demonstrate the potential of a targeted, adaptive digital therapy tool in improving language production for individuals with aphasia.

The eCALAP-rehabilitation program offers significant advantages in providing targeted rehabilitation, addressing the linguistic components most affected in each patient. Unlike many traditional therapies (Brady et al., 2016), which may rely on a one-size-fits-all approach, eCALAP is designed to tailor rehabilitation tasks based on the initial assessment and the participants progress throughout the intervention. This flexibility is particularly valuable for aphasia rehabilitation, where impairments may vary from patient to patient, affecting comprehension and/or production, words and/or sentences, with varying degrees of severity, making individualized treatment essential.

It is worth noting that P2 demonstrated more mild progress compared to the other participants. This discrepancy could be attributed to several factors. First, P2 had a longer time since the stroke (26 months), which may have influenced the brain’s plasticity and its ability to benefit from rehabilitation (Johansson, 2011). Second, aphasia severity may have also contributed to the mixed results: P2’s aphasia was the most severe, and almost every language component was severely impaired. Aphasia severity can impact negatively on recovery (The REhabilitation and recovery of peopLE with Aphasia after StrokE (RELEASE) Collaborators et al., 2022).

This study also highlights the feasibility and acceptability of using digital platforms for aphasia rehabilitation. None of the participants dropped out of the study. All the participants completed self-rehabilitation sessions between the sessions with the therapist.

Another advantage of using digital tools such as the eCALAP-rehabilitation program, is the potential to increase patients’ self-esteem (Wade et al., 2003). The use of tablets can empower patients, allowing them to take a more active role in their rehabilitation. Furthermore, digital platforms can help address challenges related to access to care. They can provide valuable support for individuals who face difficulties in attending traditional in-person sessions, whether due to geographic, financial or mobility constraints, which are common in PWA (Schmeler et al., 2009). The flexibility of self-rehabilitation sessions using the eCALAP-rehabilitation program thus not only promotes linguistic recovery but also supports patients in managing their condition independently, reducing autonomy loss, and potentially improving both their functional and emotional well-being.

Several limitations should be considered. First, only one repeated measure was used (picture naming) limiting the scope of the linguistic abilities tested. While picture naming is a valuable task for evaluating expressive language abilities, future studies could benefit from adding additional measures that assess a broader range of linguistic components, such as word repetition or sentence comprehension. However, in the context of SCED methodology, where frequent assessments are needed, it is important to find a balance between the number of measures and the feasibility of administering them multiple times a week. Furthermore, there was no control over the time spent on self-rehabilitation tasks, which have varied between participants. Future studies should ensure more consistent tracking of this variable to better understand its impact on outcomes.

Given the positive results observed in this pilot study, future research will explore replicating the study with subacute patients and include additional measures of comprehension and repetition. This will allow for a more comprehensive evaluation of the eCALAP tool’s effectiveness in addressing a wider range of aphasia symptoms.

In conclusion, although more research is needed to replicate these findings, this study suggests that eCALAP-rehabilitation could be a valuable tool for improving language performance in individuals with aphasia, offering patients an accessible, tailored, effective and cost-efficient therapy.

## Data Availability

All data produced in the present study are available upon reasonable request to the authors.

## Acknowledgments

None.

## Sources of Funding

This work was supported by: ANR-17-EURE-0017; ANR-10-IDEX-0001-02; ARS: 2023_IO_105_eCALAP; Université Paris-Est Creteil

## Disclosure

None.

## Supplementary

**Table S1:**
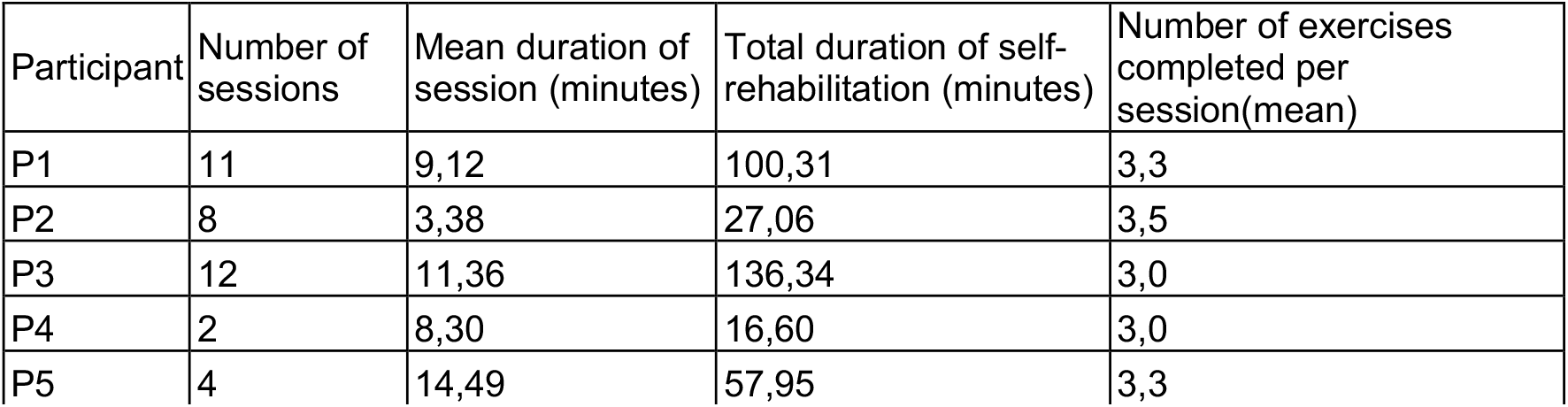
Demographics and clinical performance of the participants at baseline. F: Female; M: Male.

**Table S2:**
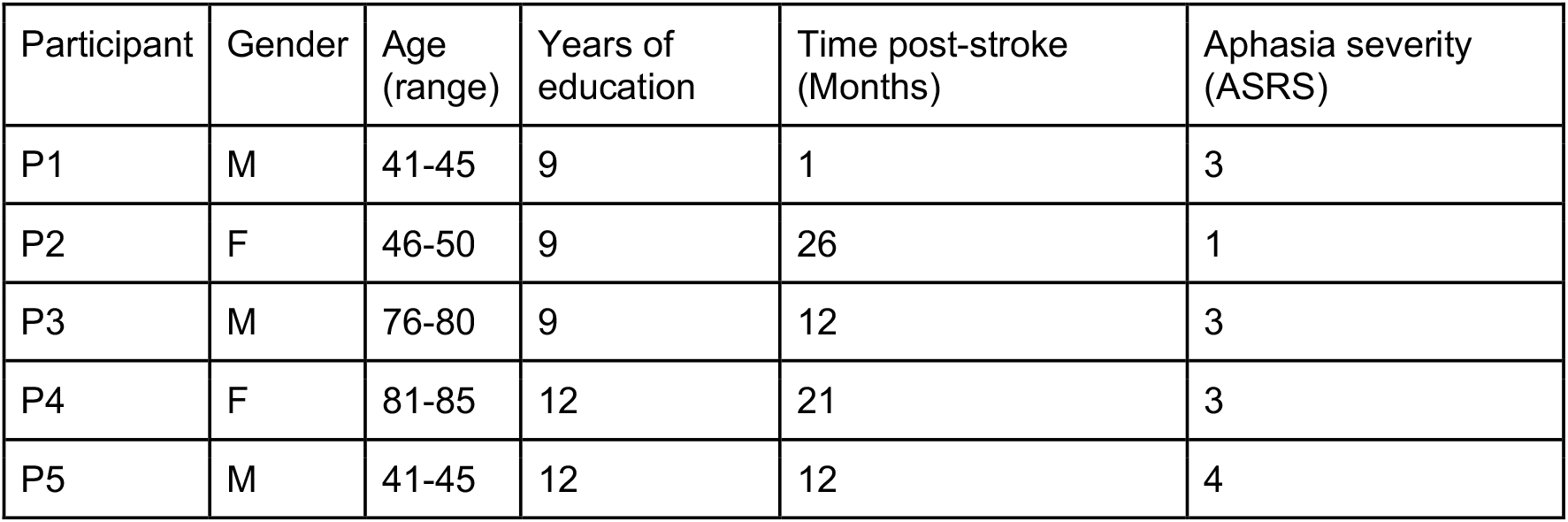
Self-rehabilitation data for each participant.

## Notes

### Competing Interest Statement

The authors have declared no competing interest.

### Clinical Trial

NCT06817642

### Funding Statement

This study was funded by ANR-17-EURE-0017; ANR-10-IDEX-0001-02; l'Agence Regionale de Sante: 2023_IO_105_eCALAP; Universite Paris-Est Creteil

### Author Declarations

Ethics committee/IRB of Universite Paris-Cite gave ethical approval for this work (IRB: 00012024-71) Ethics committee/IRB of Assistance Publique des Hopitaux de Paris gave ethical approval for this work (IDRCB: 2022-A00489-34)

## References

Bachoud-Lévi, A.-C., Dormeuil, A., & Jacquemot, C. (2022). Improving efficacy of aphasia rehabilitation by using Core Assessment of Language Processing. Annals of Physical and Rehabilitation Medicine, 65(6), 101630. 10.1016/j.rehab.2022.101630

Brady, M. C., Kelly, H., Godwin, J., Enderby, P., & Campbell, P. (2016). Speech and language therapy for aphasia following stroke. Cochrane Database of Systematic Reviews, 2016(6). 10.1002/14651858.CD000425.pub4

Bullier, B., Cassoudesalle, H., Villain, M., Cogné, M., Mollo, C., De Gabory, I., Dehail, P., Joseph, P.-A., Sibon, I., & Glize, B. (2020). New factors that affect quality of life in patients with aphasia. Annals of Physical and Rehabilitation Medicine, 63(1), 33– 37. 10.1016/j.rehab.2019.06.015

Burfein, P., Roxbury, T., Doig, E. J., McSween, M.-P., De Silva, N., & Copland, D. A. (2024). Return to work for stroke survivors with aphasia: A quantitative scoping review. Neuropsychological Rehabilitation, 1–35. 10.1080/09602011.2024.2381874

Cuperus, P., De Kok, D., De Aguiar, V., & Nickels, L. (2024). Aphasia therapy software: An investigation of the research literature and the challenges of software development. Aphasiology, 1–32. 10.1080/02687038.2024.2384542

Engelter, S. T., Gostynski, M., Papa, S., Frei, M., Born, C., Ajdacic-Gross, V., Gutzwiller, F., & Lyrer, P. A. (2006). Epidemiology of Aphasia Attributable to First Ischemic Stroke: Incidence, Severity, Fluency, Etiology, and Thrombolysis. Stroke, 37(6), 1379–1384. 10.1161/01.STR.0000221815.64093.8c

Jacquemot, C., Dupoux, E., Robotham, L., & Bachoud-Lévi, A.-C. (2012). Specificity in Rehabilitation of Word Production: A Meta-Analysis and a Case Study. Behavioural Neurology, 25(2), 73–101. 10.1155/2012/418920

Johansson, B. B. (2011). Current trends in stroke rehabilitation. A review with focus on brain plasticity: Stroke rehabilitation. Acta Neurologica Scandinavica, 123(3), 147–159. 10.1111/j.1600-0404.2010.01417.x

Krasny-Pacini, A., & Evans, J. (2018). Single-case experimental designs to assess intervention effectiveness in rehabilitation: A practical guide. Annals of Physical and Rehabilitation Medicine, 61(3), 164–179. 10.1016/j.rehab.2017.12.002

Manolov R, Moeyaert M. Recommendations for Choosing Single-Case Data Analytical Techniques. Behav Ther. 2017 Jan;48(1):97–114. doi: 10.1016/j.beth.2016.04.008. Epub 2016 May 16. PMID: 28077224.

Manolov, R., & Solanas, A. (2018). Quantifying differences between conditions in single-case designs: Possible analysis and meta-analysis. Developmental Neurorehabilitation, 21(4), 238– 252. 10.3109/17518423.2015.1100688

Parker RI, Vannest KJ, Davis JL, Sauber SB. Combining nonoverlap and trend for single-case research: Tau-U. Behav Ther. 2011 Jun;42(2):284–99. doi: 10.1016/j.beth.2010.08.006. Epub 2011 Feb 3. PMID: 21496513.

Repetto, C., Paolillo, M. P., Tuena, C., Bellinzona, F., & Riva, G. (2021). Innovative technology-based interventions in aphasia rehabilitation: A systematic review. Aphasiology, 35(12), 1623–1646. 10.1080/02687038.2020.1819957

Riley, E. A. (2017). Patient Fatigue During Aphasia Treatment: A Survey of Speech-Language Pathologists. Communication Disorders Quarterly, 38(3), 143–153. 10.1177/1525740116656330

Schmeler, M. R., Schein, R. M., McCue, M., & Betz, K. (2009). Telerehabilitation Clinical and Vocational Applications for Assistive Technology: Research, Opportunities, and Challenges. International Journal of Telerehabilitation, 59–72. 10.5195/ijt.2009.6014

Shrubsole, K., Worrall, L., Power, E., & O’Connor, D. A. (2018). Priorities for Closing the Evidence-Practice Gaps in Poststroke Aphasia Rehabilitation: A Scoping Review. Archives of Physical Medicine and Rehabilitation, 99(7), 1413-1423.e24. 10.1016/j.apmr.2017.08.474

Tarlow, K. R., & Penland, A. (2016). Outcome assessment and inference with the percentage of nonoverlapping data (PND) single-case statistic. Practice Innovations, 1(4), 221–233. 10.1037/pri0000029

Tate, R. L., & Perdices, M. (2019). Single-Case Experimental Designs for Clinical Research and Neurorehabilitation Settings: Planning, Conduct, Analysis and Reporting (1^re^ éd.). Routledge. 10.4324/9780429488184

The REhabilitation and recovery of peopLE with Aphasia after StrokE (RELEASE) Collaborators, Brady, M.C., Ali, M., VandenBerg, K., Williams, L. J., Williams, L. R., Abo, M., Becker, F., Bowen, A., Brandenburg, C., Breitenstein, C., Bruehl, S., Copland, D. A., Cranfill, T. B., Di Pietro-Bachmann, M., Enderby, P., Fillingham, J., Galli, F. L., Gandolfi, M., … Wright, H. H. (2022). Dosage, Intensity, and Frequency of Language Therapy for Aphasia: A Systematic Review–Based, Individual Participant Data Network Meta-Analysis. Stroke, 53(3), 956–967. 10.1161/STROKEAHA.121.035216

Vaezipour, A., Campbell, J., Theodoros, D., & Russell, T. (2020). Mobile Apps for Speech-Language Therapy in Adults With Communication Disorders: Review of Content and Quality. JMIR mHealth and uHealth, 8(10), e18858. 10.2196/18858

Wade, J., Mortley, J., & Enderby, P. (2003). Talk about IT: Views of people with aphasia and their partners on receiving remotely monitored computer-based word finding therapy. Aphasiology, 17(11), 1031–1056. 10.1080/02687030344000373

